# Segment and Slice: A Two-Step Deep Learning Pipeline for Opportunistic Vertebral Fracture Detection in Computed Tomography

**DOI:** 10.1101/2022.11.26.22282267

**Authors:** Carl G. Glessgen, Joshy Cyriac, Shan Yang, Sebastian Manneck, Hildegard Wichtmann, Jakob Wasserthal, Balazs K. Kovacs, Dorothee Harder

**Affiliations:** Clinic of Radiology and Nuclear Medicine, University Hospital Basel, University of Basel, Basel, Switzerland; Division of Radiology, Geneva University Hospitals, Rue Gabrielle-Perret-Gentil 4, 1205, Geneva, Switzerland

**Keywords:** computed tomography, vertebral segmentation, fracture detection, deep learning, neural networks

## Abstract

**Objectives:** An end-to-end method is introduced to build a combined segmentation-classification pipeline using deep learning for opportunistic fracture detection in CT spine images of varying field-of-views.

**Materials and Methods:** This retrospective study builds on 452 CTs of the lumbar/thoracolumbar spine. Patients were included based on the evidence of ≥ 1 vertebral body fracture and excluded in case of history of spinal surgery or pathologic fractures. The collective was split into training/validation (405) and test (47) sets. An open-source pre-segmented spine dataset was used to train a preliminary segmentation model, which was applied on the training set. The resulting segmentation was post-processed to remove posterior vertebral structures and if needed manually refined by a radiologist. Using the refined version as new training data, a final segmentation nnU-net was trained. Sagittal slices from each vertebra were labelled individually with regard to fracture evidence. Slices without signs of fracture were used as negative class. 27,019 slices (20,396 negative, 6,623 positive) trained a classification algorithm using resnet18. Two senior readers independently assessed fractures in the test set to obtain a consensual ground truth. The segmentation-classification pipeline was applied to the test set and compared to the ground truth.

**Results:** The segmentation model correctly segmented 330/339 (97%) vertebrae. Considering every segmented vertebra, the classifier detected fractures with 88% sensitivity, 95% specificity and 93% accuracy.

**Conclusion:** Our two-step method can help to detect spine fractures on images of varying field-of-views, with an accuracy comparable to that of a radiologist in-training. The final models as well as our code material are available at https://github.com/usb-radiology/VertebraeFx.

## Introduction

For a radiologist, accurately reporting and classifying vertebral fractures is an essential skill since they belong to the routine workflow of almost any health system. De facto, the prevalence of any kind of vertebral deformity was estimated at 25% in a large cohort of women over 50 years ^1^ and a recent meta-analysis yielded a regional range of up to 26% for vertebral fracture prevalence ^2^. As a result, vertebral fractures account for 5% of all direct costs for care of osteoporosis and for up to 21% of the societal aftermath of osteoporosis ^3,4^. It is all the more reason to report them carefully since the risk of a new vertebral fracture increases up to 5-fold in the year following an incident vertebral fracture ^5^.

Nevertheless, the radiologic identification of vertebral fractures is still insufficient. In a large multicentric study, the underdiagnosis of vertebral fractures on radiographs ranged between 29.5% and 45% ^6^, while a meta-analysis of studies assessing vertebral fractures estimated their mean reporting rate at 27% ^7^. Radiological reports for long-term hospital patients are particularly at risk of lacking a comprehensive description of vertebral fractures ^8^. The constantly rising use of diagnostic imaging ^9^ makes correcting this deficiency even more challenging, as radiology teams have to cope with a sustained increase in imaging volumes.

At the same time, research in the field of automatic fracture detection and spine segmentation of CT images has seen an exponential growth over the years, as have the technological solutions available. Specifically, solutions for fracture detection shifted from the use of machine learning ^10–14^ to deep learning models trained on 2D ^15,16^ or more recently 3D images ^17,18^. Implementing those technologies and ideas appears as a credible way to standardize fracture reporting and to support radiologists in their real-life clinical workflow. On the other hand, implementing the work of others is limited by software compatibility, variability in CT images from different scanners, and the understanding of the technology offered. Also, many models cited above do not report the ability to process field-of-views of varying size.

Our goal was to contribute to this growing research field and to develop an opportunistic fracture detection model by leveraging deep learning technology. The network should be able to process CT images of varying field-of-views and accessible to other research teams via the open-source tools presented. We intend to evaluate the method as proof-of-concept, to pinpoint its limitations and identify future improvements.

## Material and Methods

The institutional review board waived informed consent.

### Study population

This study is retrospective and builds upon CT studies of the lumbar and thoracolumbar spine performed between January 2015 and December 2020 at our institution. The collection of datasets was performed using our department’s radiology report crawler. The sole inclusion criterion was any mention of at least one vertebral body fracture in the report. Cases were excluded based on the following criteria: i) history of spinal fusion, vertebral augmentation or extensive spinal surgery, ii) concurrent or past spondylodiscitis, iii) presence of vertebral metastases or malignant lesions including pathologic fractures, iv) osteolysis arising from adjacent malignant tissue and v) studies performed for myelography. Fractures were classified using the AO spine classification^19^ which is being used at our institution.

The study collective was split into training and test sets with a 9:1 ratio. The cases for the validation set were randomly selected from the training set included in the study.

### Imaging

All included studies were non-enhanced and acquired by one of three multi-detector CT scanners (Somatom Definition Flash, Somatom Definition Edge, Somatom Force) from Siemens (Siemens Healthineers, Erlangen, Germany). The acquisition parameters are listed in Table 1. For each case, the sagittal reconstruction using a bone kernel was extracted from the PACS (GE Healthcare, Chicago, USA). The web-based framework for image analysis Nora (Nora Medical Imaging Platform Project, Freiburg im Breisgau, Germany) was used for organisation, visualisation and processing of images.

**Table 1.**
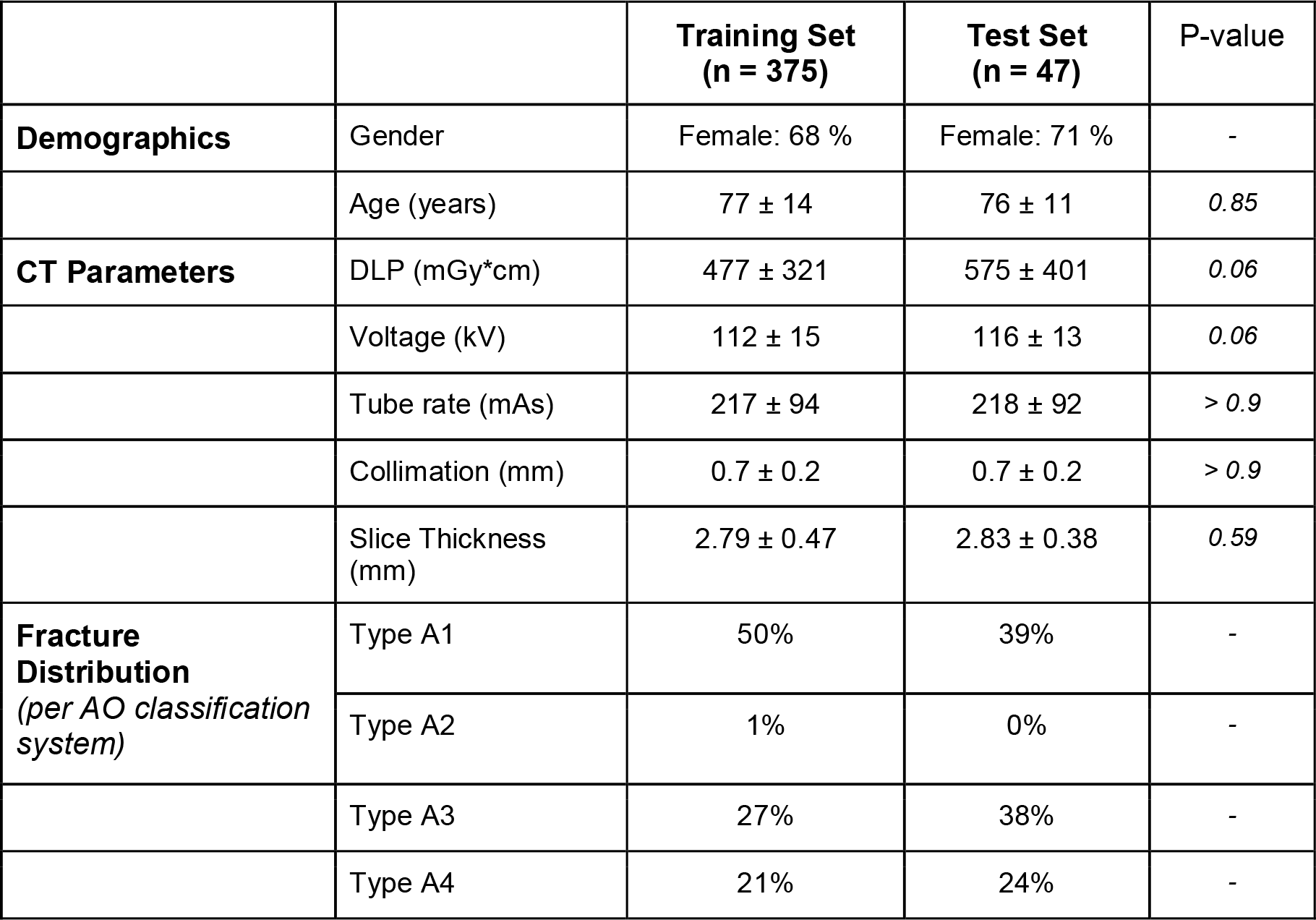
Overview of the main demographic and CT acquisition parameters in the training and validation sets. Values are presented as mean ± standard deviation.

|  |  | **Training Set**<br>**(n = 375)** | **Test Set**<br>**(n = 47)** | P-value |
| --- | --- | --- | --- | --- |
| **Demographics** | Gender | Female: 68 % | Female: 71 % | - |
|  | Age (years) | 77 ± 14 | 76 ± 11 | 0.85 |
| **CT Parameters** | DLP (mGy*cm) | 477 ± 321 | 575 ± 401 | 0.06 |
|  | Voltage (kV) | 112 ± 15 | 116 ± 13 | 0.06 |
|  | Tube rate (mAs) | 217 ± 94 | 218 ± 92 | > 0.9 |
|  | Collimation (mm) | 0.7 ± 0.2 | 0.7 ± 0.2 | > 0.9 |
|  | Slice Thickness (mm) | 2.79 ± 0.47 | 2.83 ± 0.38 | 0.59 |
| **Fracture Distribution**<br>(per AO classification system) | Type A1 | 50% | 39% | - |
|  | Type A2 | 1% | 0% | - |
|  | Type A3 | 27% | 38% | - |
|  | Type A4 | 21% | 24% | - |

### System overview

The study follows a two-step approach by training a vertebral segmentation model and a fracture classification model independently. Studies acquired during the daily workflow and corresponding to a specific label (i.e. “CT of the spine”) can be automatically submitted from the PACS to the segmentation-classification pipeline built in the Nora framework. Following segmentation, the segmented vertebral volumes are presented as series of 2D-slices to the classification algorithm. If the classification model detects a fracture in one of the slices, the vertebra at stake is considered affected by a fracture. Consecutively, a report is sent back to the PACS with a listing of all explored vertebrae in the field-of-view and their status regarding fracture.

### Vertebral segmentation algorithm

A preliminary model was trained with the open-source vertebral segmentation dataset of VerSe2019 ^20^ consisting of pre-segmented healthy spines, using a nnU-net ^21^ which is described in the framework’s source paper. All cases from the training set were segmented by this preliminary model. In a second step, automated image post-processing techniques such as erosion, dilation and connected components were used to remove the posterior vertebral structures from the segmentation mask, to split segmentation masks possibly covering multiple vertebrae and to recount the vertebral order in cranio-caudal orientation starting from L5 upwards, Figure 1. Following segmentation and post-processing, cases were manually refined, if needed, by C.G.G. (resident in-training with 4 years of experience), using Nora framework’s built-in tools.

**Figure 1.**
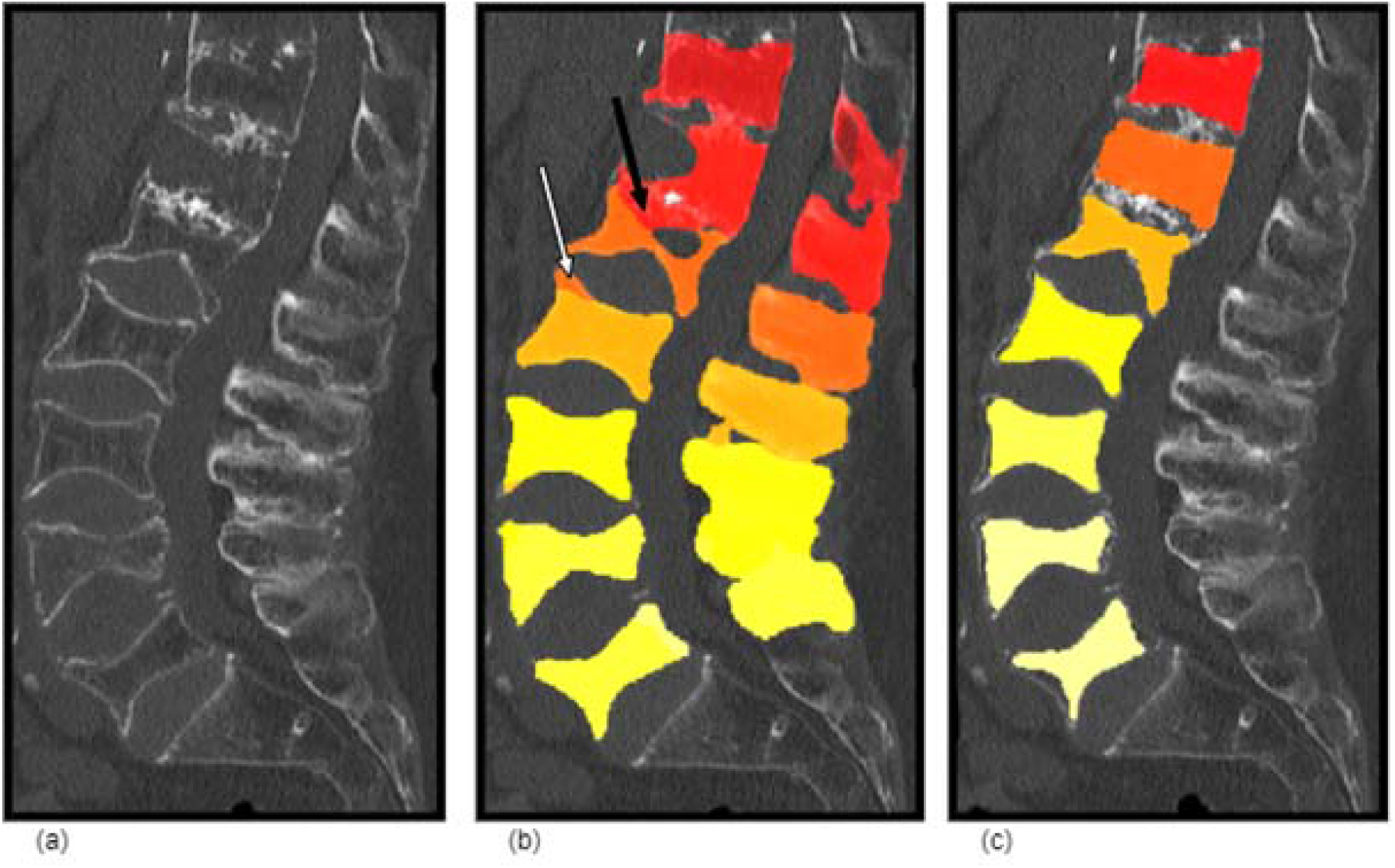
Overview of a typical segmentation inconsistency in a male patient in his 80s with a history of low energy fall and back pain. Note the heavy degeneration of the spine and partial destruction of Th12 (a). Raw segmentation mask obtained from the open-source data and the preliminary segmentation model (b). The anterior upper endplate of L2 (white arrow) has been wrongly mapped and the intervertebral disk between Th12-L1 (black arrow) has been segmented because of blurry calcifications within it. After correction, the posterior structures have been removed and Th12 is correctly demarcated (c). The last mask was used as ground truth to train the final nnU-net.

Finally, the segmented data was used to train a nnU-net as the final segmentation model. This model served as entry point of the segmentation-classification pipeline and was later applied onto the test set to extract the vertebral volumes to classify.

### Fracture classification algorithm

Employing the accurately segmented training cases, the content of each vertebral volume was extracted as batches of sagittal slices with an in-plane resolution of 2 mm to 3 mm, Figure 2. The 5 outer slices on each side of a vertebral volume were removed to maintain homogeneity throughout the data. By “shaving” the sides of the vertebrae off, their often heavily degenerated edges are removed and the vertebral slices remaining are better suited for fracture identification.

**Figure 2.**
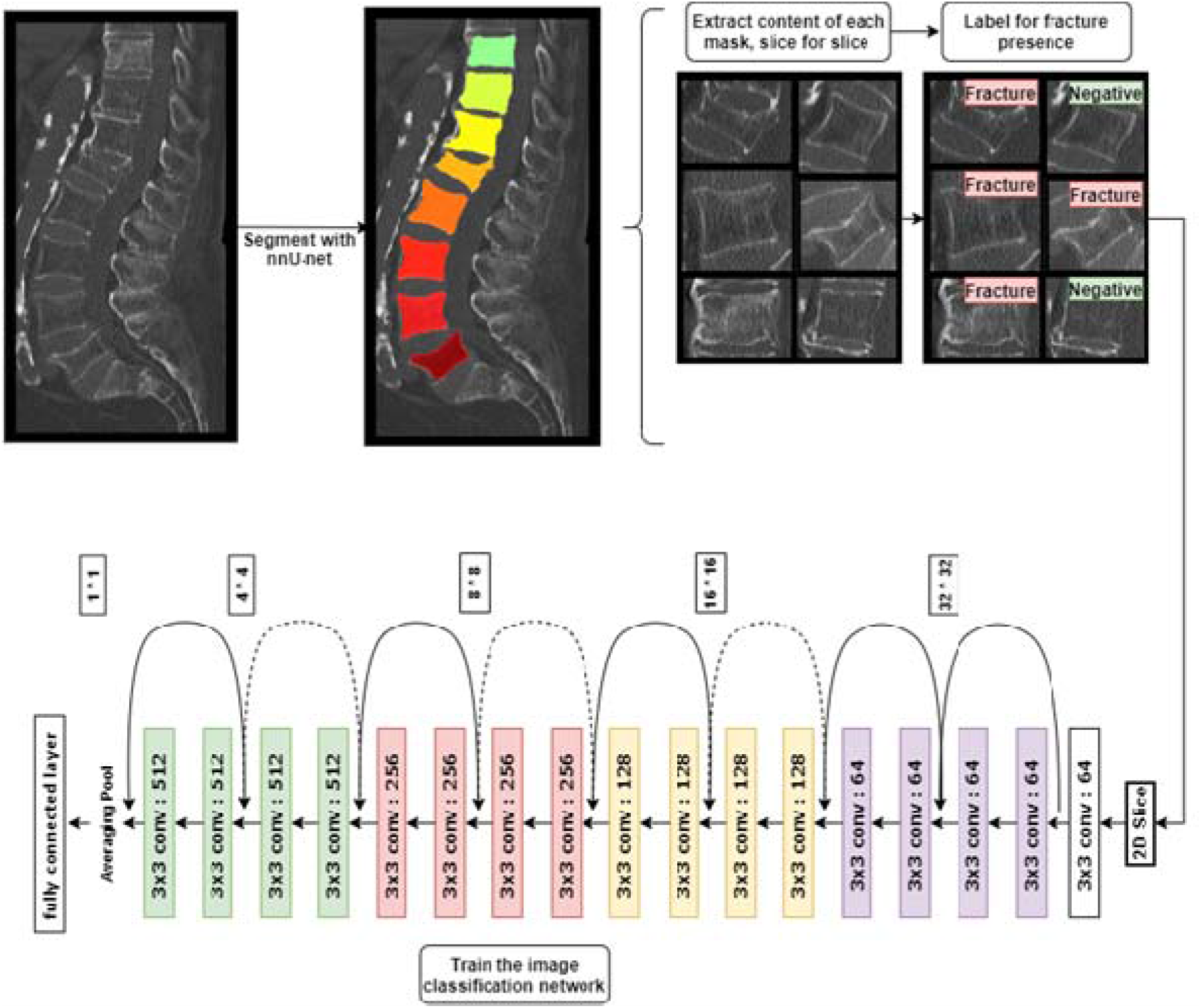
Overview of the training process. Case is a female patient in her 70s with chronic back pain. The raw data was segmented using nnU-net. The content of each vertebral mask (batches of 2D-images) was extracted and labelled at the slice level. Every slice generated was then used for training of the classification network. The architecture presented is the 18-layer version of ResNet.

For each vertebra in the training set, slices showing fracture stigmata as well as the fracture grade were manually recorded by C.G.G. Each study’s report was used as reference to define fractures. In the event of missing or ambiguous report information, unclear cases were read in consensus by C.G.G and D.H. (fellow-trained musculoskeletal radiologist with 8 years of experience). Relying on the slice coordinates recorded, each slice inside a vertebral volume was assigned a label depending on fracture presence (i.e. “fracture” or “negative”). Negative cases were provided by the spared vertebrae inside each dataset, as well as by the slices showing no fracture stigmata inside one injured vertebra.

A classification algorithm was trained using resnet18 ^22^. The batch size was set to 32 and optimized with Stochastic Gradient Descent (SGD) with an initial learning rate of 0.001 and a momentum of 0.9. The learning rate was reduced every 7 epochs by a gamma factor of 0.1. For data augmentation the following transformations were applied to the images during training: intensity normalization, random rotation with a rotation angle of [-□/12,/12] radians, random horizontal flipping, random zoom with factor of [0.9,1.1] and as a final step a resize transformation with either padding or cropping to size 256 × 256. The model was implemented using Pytorch-lightning ^23^ and TIMM ^24^. Both the final segmentation and classification pipeline are available at https://github.com/usb-radiology/VertebraeFx.

### Assessing the test set

Two senior readers, D.H. and S.M. (fellow-trained musculoskeletal radiologist with 2 years of experience), blindly and independently assessed the cases within the test set with regard to fractures and fracture grade (per AO classification system). The reading was performed on standard workstations using Centricity’s PACS (GE Healthcare). Differences in the reading scores were read in consensus 4 weeks after the initial reading to produce an absolute ground truth. Two readers in-training, C.G.G and H.W (1.5 years of experience), read the same test cases by listing the fractures they found and graded them using the AO scale.

The algorithmic pipeline (segmentation and classification models) were applied to the test set. The segmentation model segmented each vertebra and located them anatomically by counting upwards from L5. Second, the classification model extracted the slices inside the vertebrae and classified each of them. A fracture was defined if at least one slice within the vertebra was classified as having a fracture. The results were compared to the absolute ground truth as well as to the scores of both readers in-training.

### Statistical Analysis

All available cases fitting the inclusion criteria were included in the study. Cohen’s Kappa score was used to assess the interrater agreement over the test set. A p-value of 0.05 was used to infer statistical significance from the demographic and CT acquisition parameters, using independent T-tests, Table 1.

The evaluation of the classification model’s performance compared to the ground truth is quantified through usual measures of diagnostic accuracy, Table 2. Statistical analysis was performed using IBM SPSS Statistics in combination with statistic libraries of the Python framework.

**Table 2.**
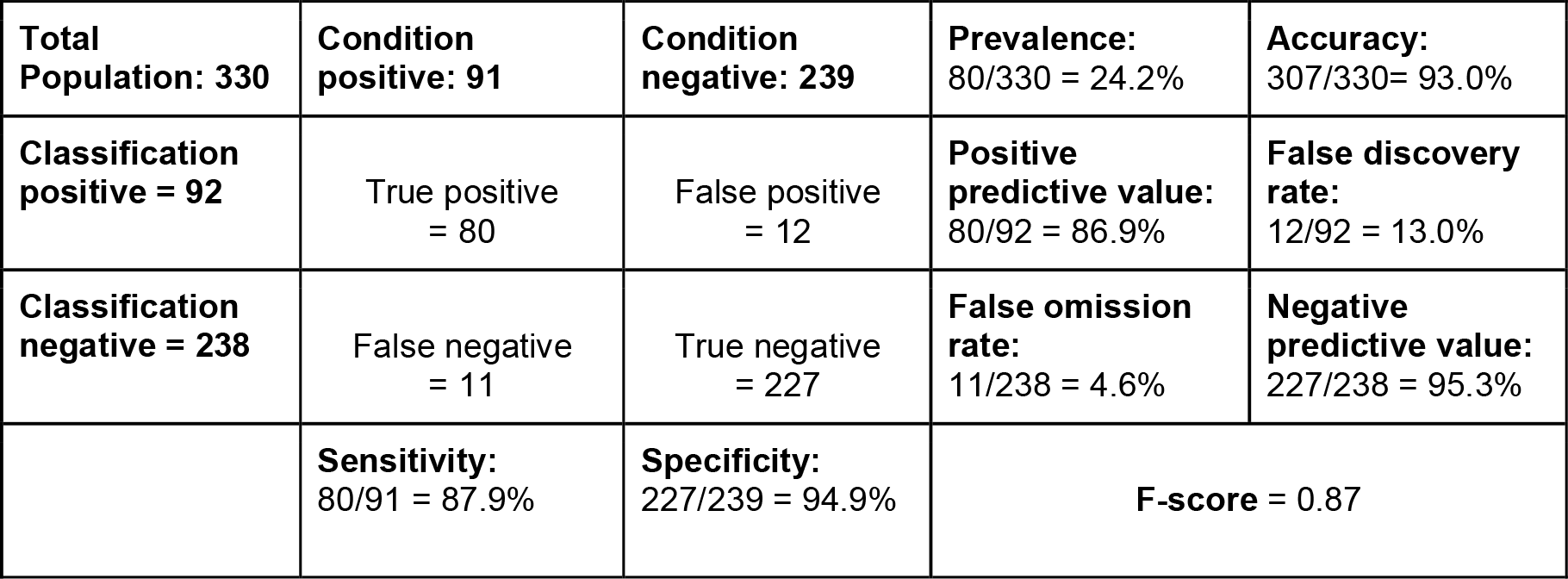
Confusion matrix of the classification model after processing the correctly segmented vertebrae, compared to the test set’s consensual ground truth.

|  |  |  |  |  |
| --- | --- | --- | --- | --- |
| **Total Population: 330** | **Condition positive: 91** | **Condition negative: 239** | **Prevalence:**<br>80/330 = 24.2% | **Accuracy:**<br>307/330= 93.0% |
| **Classification positive = 92** | True positive<br>= 80 | False positive<br>= 12 | **Positive predictive value:**<br>80/92 = 86.9% | **False discovery rate:**<br>12/92 = 13.0% |
| **Classification negative = 238** | False negative<br>= 11 | True negative<br>= 227 | **False omission rate:**<br>11/238 = 4.6% | **Negative predictive value:**<br>227/238 = 95.3% |
|  | **Sensitivity:**<br>80/91 = 87.9% | **Specificity:**<br>227/239 = 94.9% | **F-score = 0.87** |  |

## Results

### Study population

The study collective consisted of 452 cases [308 women, mean age 79 ± 12 years and 244 men, mean age 72 ± 16 years] which were randomly split into a training set (405 cases) and a test set (47 cases), Figure 3. There were no differences in age and gender distribution between the cases in the validation and training sets, p > 0.05, Table 1. There were more wedge-compression fractures in the training set (50%) than in the test set (39%). The distribution of other fracture types between both sets remained fit for comparison.

**Figure 3.**
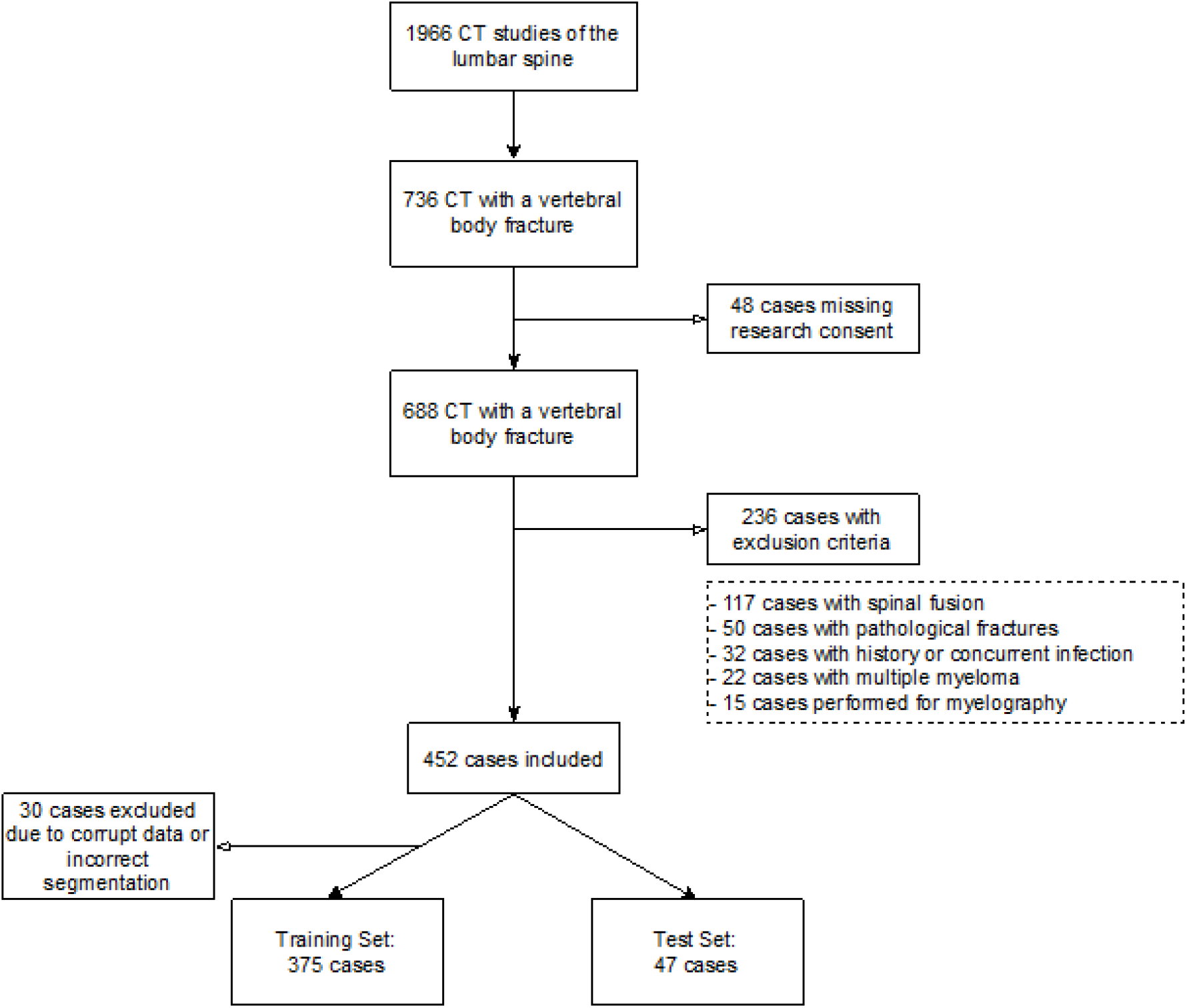
Flowchart outlining the process of patient inclusion in the study collective.

### Imaging

CT studies were heterogeneous in scan range, from tailored field-of-views with four vertebrae depicted up to thoracolumbar protocols with twelve vertebrae inside the field-of-view. Some studies did not show the lowest lumbar vertebrae (e.g. L4 or L5). The acquisition parameters are listed in Table 1. The matrix sizes ranged from 608 × 512 to 2228 × 1024 pixels, Figure 4.

**Figure 4.**
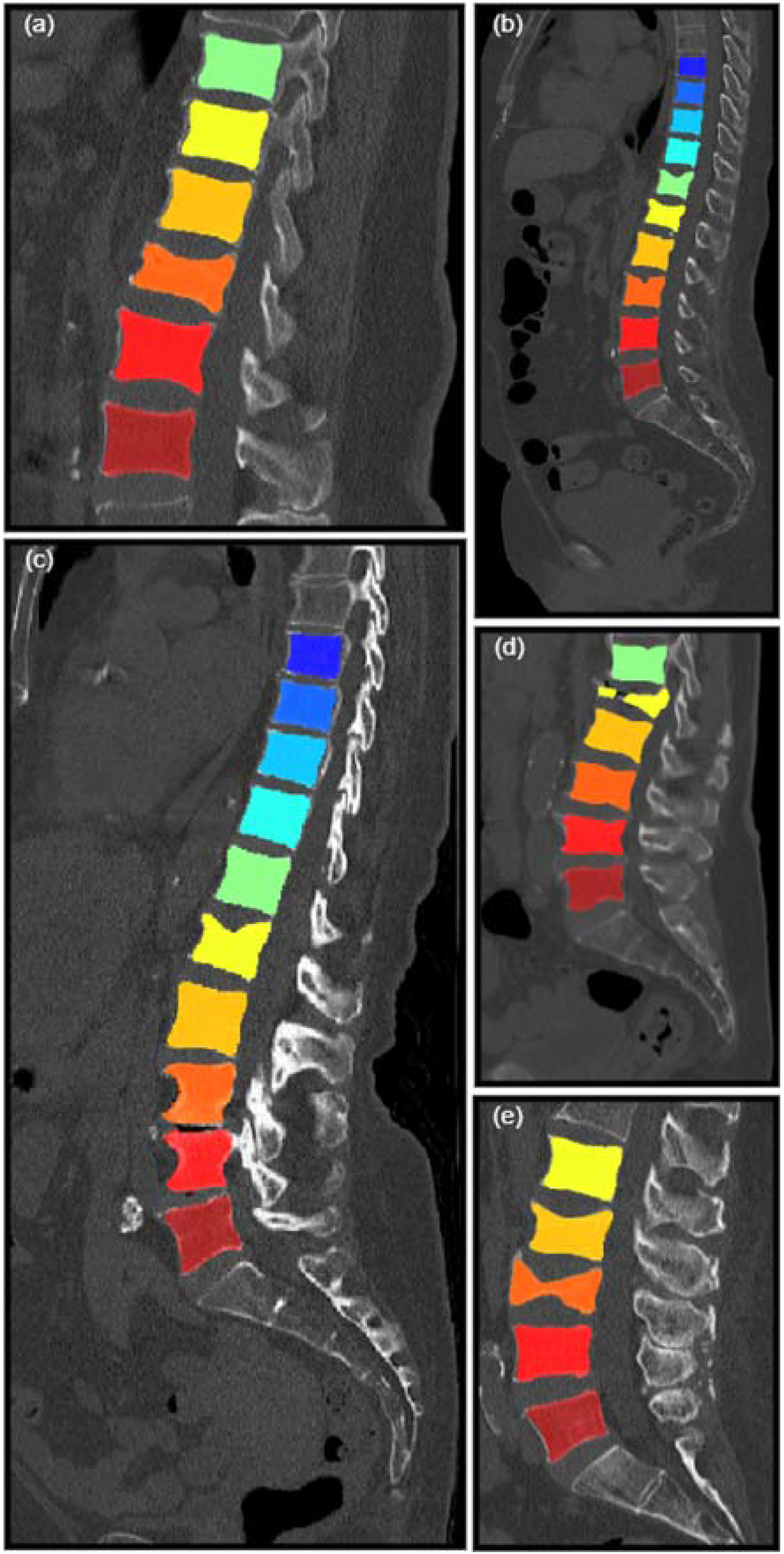
Different cases from the test set after segmentation, with varying matrix sizes: (a) 658 × 512, (b) and (c) 1046 × 512, (d) 797 × 512 and (e) 777 × 512. Note the excessive removal of the most cranial vertebrae in cases (b) and (c). Case (a) is a typical example of a shifted vertebral atlas, due to the lowest vertebra not being L5. However, all vertebrae are correctly segmented.

### Vertebral segmentation algorithm

The inconsistencies of the preliminary segmentation via the data of VerSe2019 included errors due to anatomical variations (e.g. a 6th lumbar vertebra), inadequate coverage or numeration of a vertebra (Figure 1) and fused masks over two vertebrae. In total, 30/405 cases could not be segmented or post-processed due to major spine degeneration, anatomical variations unworkable by the post-processing step, corrupt data or data too large to process. This resulted in a final collective of 375 segmented cases for the training set. Following preliminary segmentation and post-processing, 67/375 cases had to be corrected manually by C.G.G.

### Fracture classification algorithm

There were 438 fractures recorded in the training set’s 375 cases. AO classification was used. 220 fractures (50%) were type A1, 5 (1%) were type A2, 119 (27%) were type A3, 94 (21%) were type A4. After segmenting every vertebra and extracting every labeled slice, a collection of 27,019 2D-pictures was obtained: 20,396 of the negative class, 6,623 of the positive class. The model was trained up to 5 epochs.

### Test set

The test set consisted of 47 cases encompassing a total of 339 vertebrae. The senior readers D.H. and S.M. reported 87 and 95 fractures, respectively, with 11 discrepant cases out of the 339. The corresponding interrater agreement was of 0.86 (Cohen’s Kappa Score). After consensus reading, the ground truth was set at 91 fractures. 35 fractures (38%) were type A1, 34 (37%) were type A3, and 22 (24%) were type A4.

The in-training readers C.G.G. and H.W. reported 82 and 92 fractures, respectively, with 10 discrepant cases out of the 339. The corresponding interrater agreement was of 0.91 (Cohen’s Kappa Score). The respective sensitivity of C.G.G. and H.W. was 82.4% and 86.8%, their specificity was 97.1% and 94.5%.

The segmentation model correctly segmented 330/339 vertebrae (97.3%). 9 vertebrae were not segmented due to the post-processing step programmed to remove vertebrae edging the field-of-view. 4 cases from the 47 were correctly segmented but not rightfully located. This was due either to the presence of an anatomical variation (6^th^ lumbar vertebra) or because some of the considered spines did not comprise an L5 (e.g., lowest was L3). Consequently, and because the segmentation started at L5, the whole vertebral sequence was shifted upwards and accurate location could not be achieved in an automated fashion. From a per-case point of view, 38/47 cases (80.8%) were correctly segmented and anatomically identified.

The classification model reported 92 fractures out of the 330 segmented vertebrae, Table 2. There were 80 true positives, 227 true negatives, 12 false positives and 11 false negatives. This matches a sensitivity of 87.9%, a specificity of 94.9%, an accuracy of 93.0%, and positive and negative predictive values of respectively 86.9% and 95.3%. From a per-case point of view, 30/47 (63.8%) cases were correctly classified. All missed fractures (11/11) were of type A1 per AO classification system, i.e. wedge-compression fractures.

False positive cases were caused by cortical irregularities, Schmorl nodes and osteochondrosis, Figure 5.

**Figure 5.**
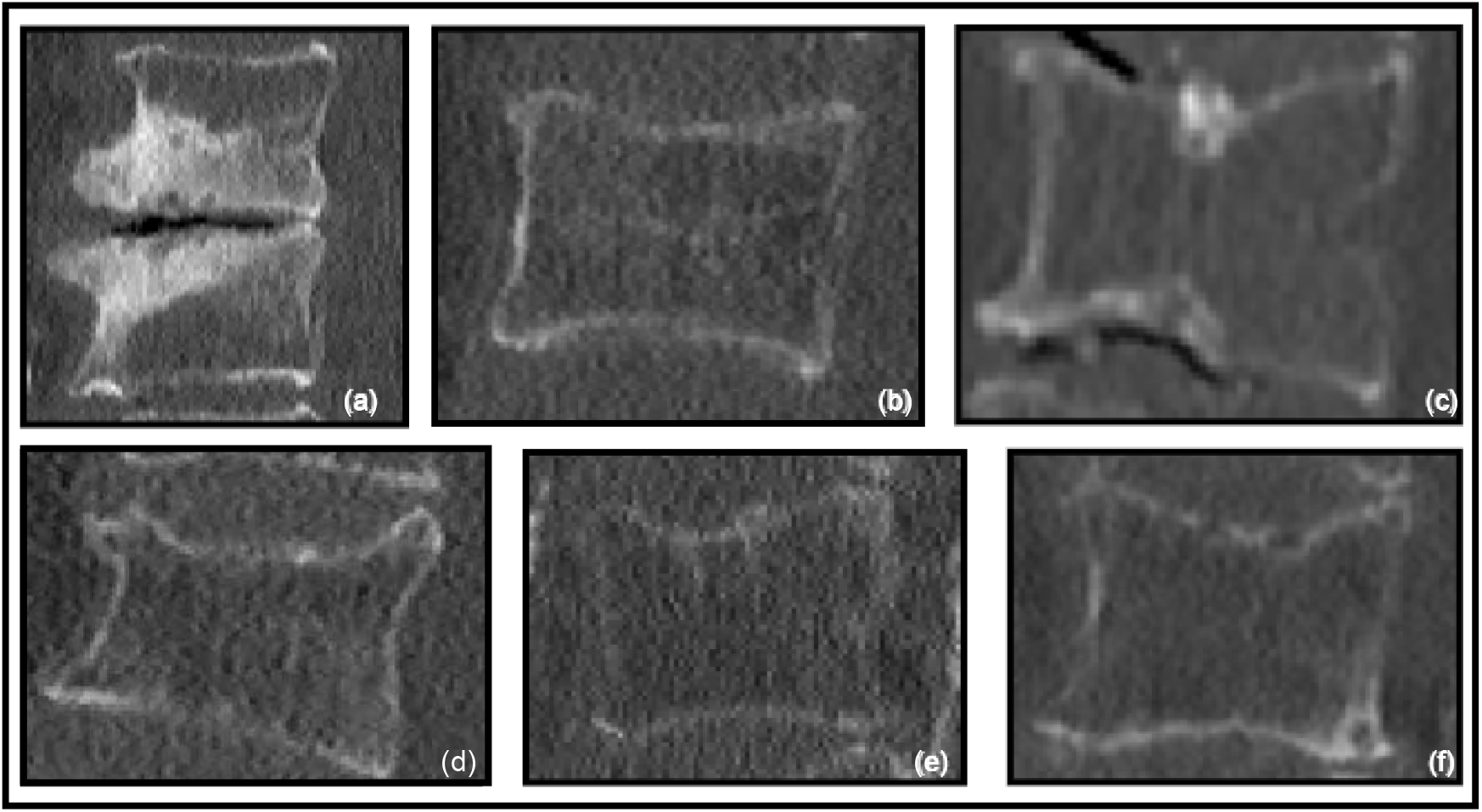
Examples of failed classifications. The upper row presents false positive cases which were due to: sclerotic endplates following osteochondritis (a), unspecific cortical irregularities (b) and calcified Schmorl nodes (c). The lower row presents false negative cases: three wedge-compression fractures (d-f), type A1 per AO classification system. Note the various image grain, resolution and vertebral density across the set.

A standard output to the PACS from the algorithmic pipeline fracture assessment is proposed, Figure 6. In addition, the weights of both segmentation and classification models as well as the image post-processing code for the segmentation pipeline are made available at github.

**Figure 6.**
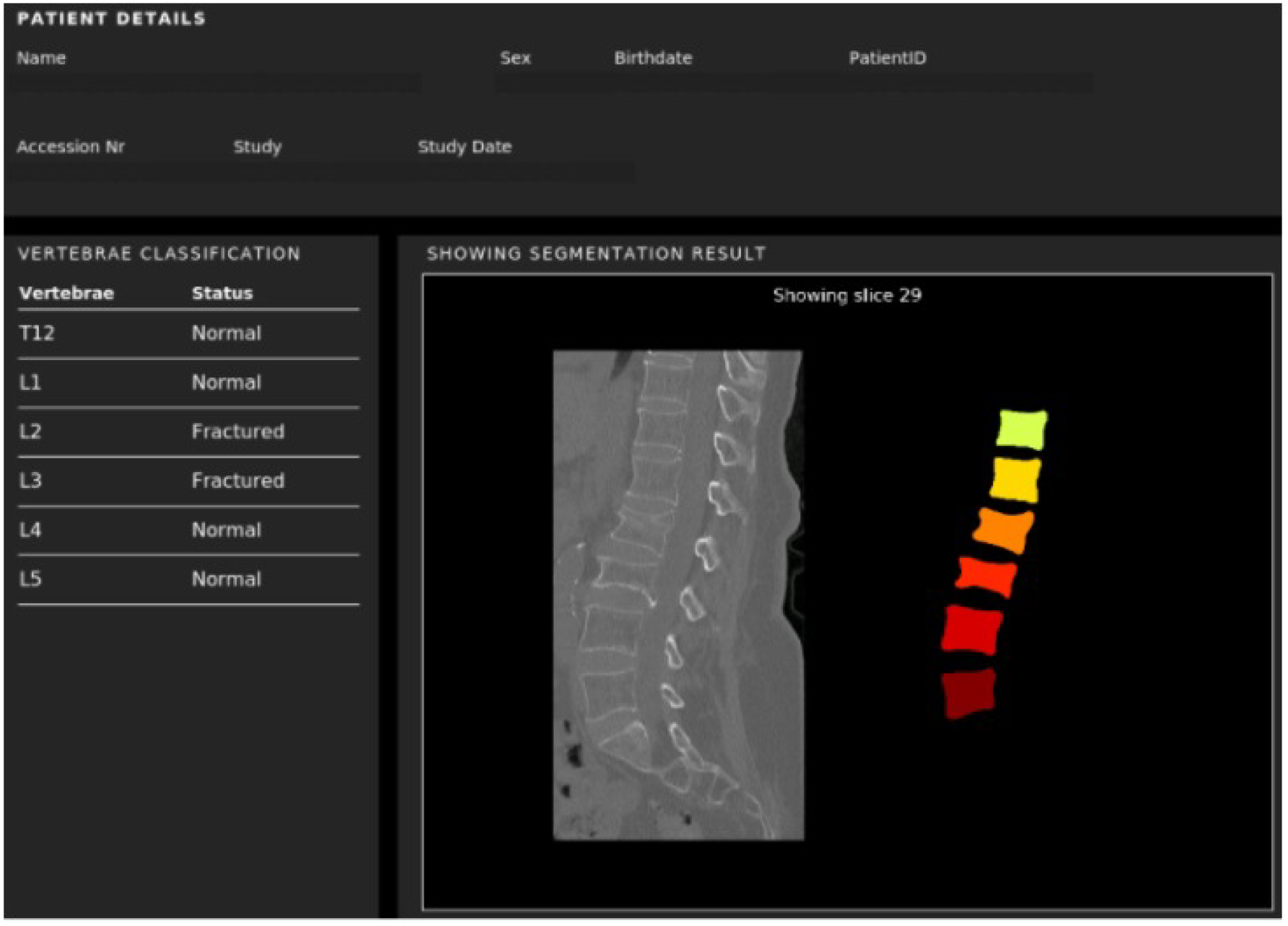
Example of a possible model output. The diagram is produced aside from the PACS (on the server on which the model is running) and can be transferred to it afterwards. Note the indication to retrace the slice presented.

## Discussion

The aim of this study was to develop an algorithmic pipeline using deep learning, to detect fractures from CT images with varying scan ranges. The results show that sensitive (88%), specific (95%) and accurate (93%) detection can be achieved by training a network over a medium-sized collective of 400 cases. In terms of diagnostic accuracy measures, our model fits the range of previously published studies (sensitivity, specificity and accuracy measures of fracture detection ranged between 81-96%, 77-94% and 89-90% respectively), ^10,11,13–18^.

The scope of techniques used previously is very broad, ranging from support vector machines ^13^ to 3D-based convolutional networks ^17,18^. The latest state-of-the-art research comes from the work of Husseini and al. ^17^ which combined an unsupervised 3D-based CNN for representational learning of vertebral shapes and a discriminative MLP (multi-layer perceptron) for classification, with a recall rate of up to 100%. Instead, our work builds on a 2D-based CNN to classify fractures from series of slices input to the network. 2D-based models are yet better understood and adjustable than 3D-models, and generally need much less GPU for training. On the other hand, the vertebra’s global assessment is better with a 3D-based approach, as valuable information can be aggregated along the z-axis. Other features than the fracture diagnostic, such as the fracture type or age, could then be predicted without requiring additional reading. It is not clear whether the amount of data needed would have to grow from the 400 presented here or the 2700 vertebrae processed by Husseini and al. Other studies have determined the fracture type in CT studies using ML ^11^, but accurate prediction of a spine fracture’s age has to our knowledge not been achieved yet.

We deliberately chose a two-step over a one-stop-shop approach because separating segmentation and classification tasks made the system more adjustable. Instead, for example, of using a Faster-RCNN ^25^ to simultaneously train an object detector (which would detect the bounding boxes for the vertebral bodies) and a classifier on those objects. In addition, valuable information such as bone density values can be extracted from the segmented volume. This kind of two-step approach is not new but the type of data input to the network can vary greatly. Tomita and al have also used CNNs ^15^ but with whole CT slices as inputs instead of smaller vertebral slices. This makes the algorithm susceptible to learning confounders in the volume instead of relying entirely on the considered vertebra. Bar and al. ^16^ used patches of vertebral slices provided by a CNN as input, and a recurrent neural network as classifier, but needed a patient collective of around 3700 cases for similar diagnostic accuracy results. In our case, slicing relatively few vertebrae into 2D-images translated into a large image database, which was sufficient for accurate training.

Regarding our work, it has to be noted that all missed cases were minor wedge-compression fractures which do not call upon surgical management. False positives were mainly due to sclerotic endplates or disc degeneration. In comparison with our in-training radiologists who run the diagnostic front during shifts, the algorithm showed a higher sensitivity than both readers and a slightly inferior specificity. Interestingly, its detection pattern (i.e. high sensitivity and consequently lower specificity) closely matched the pattern of our resident with < 2 years of experience. In terms of processing time, our segmentation-classification pipeline on the Nora platform needed on average 2-3 minutes per case, once it had received PACS images. This delay is compatible with its assigned task of opportunistic fracture detection. Past studies reported processing times from 50 min/case ^10^ to 0.02 sec/case ^15^.

Our work holds some limitations. First and foremost, we disclose an important selection bias on our study population by excluding studies with surgical material and pathological fractures for training. Obviously, the potential of the trained algorithmic pipeline if presented with such cases – especially the segmentation part of it – is not clear. The preliminary nature of this study made us focus on the flexibility and continuity of the pipeline instead of its exhaustiveness. Simultaneously, although the input data was heterogeneous, CT scanners used for this study were from one unique vendor. Both the lack of scanner and patient diversity could limit generalization of our findings. Especially the high specificity achieved by our model could deteriorate if confronted with infectious or lytic endplate changes, for example. Additional evaluation and further training of the model is needed to validate its clinical effectiveness. Second, the post-processing we applied to refine the preliminary segmentation model (nnU-net) led to inconsistencies. Mainly, it removed the most cranial vertebra, even if fully displayed. Consequently, the first cranial vertebra in some cases (9 vertebrae from 339) was neither segmented nor assessed by the classification model. Also, we excluded 30 cases (approximately 7%) from the training collective because of the impossible segmentation using the nnU-net and our post-processing code. This selection bias needs to be acknowledged when assessing the network’s diagnostic accuracy presented. Third, our model counts the vertebrae from the bottom up starting with L5. This implies that cases with anatomical variations or cases not showing L5 might be wrongly mapped. For this reason, our report displays the fracture list but also the segmentation mask and a snapshot of the spine to ensure proper identification of the injured vertebrae.

As a conclusion, it was shown that a flexible vertebral segmentation and fracture detection algorithm can be trained over a relatively modest collective size. Standardized reporting of fractures could benefit from it by systematically integrating the model’s output into the report. Further works should focus on improving the model’s sensitivity and segmentation performance, both of which are key to grant added value for the radiologist.

## Data Availability

All code data in the present study are available online at https://github.com/usbradiology/VertebraeFx.

## Notes

### Competing Interest Statement

The authors have declared no competing interest.

### Funding Statement

This study did not receive any funding

### Author Declarations

Ethics committee/IRB of the University Hospital Basel gave ethical approval for this work.

